# Using Healthcare Claims Data to Identify Health Disparities for Individuals with Diagnosed and Undiagnosed Familial Hypercholesterolemia

**DOI:** 10.1101/2024.07.26.24311087

**Authors:** Mary P. McGowan, Chao Xing, Amit Khera, Chun-Yuan Huang, Yanqiu Shao, Michelle Xing, Eric J. Brandt, Diane E. MacDougall, Catherine D. Ahmed, Katherine A. Wilemon, Zahid Ahmad

**Affiliations:** The Family Heart Foundation, Fernandina Beach, FL; Division of Cardiology, Department of Internal Medicine, Dartmouth Hitchcock Medical Center, Lebanon, NH; McDermott Center for Human Growth and Development, University of Texas Southwestern, Dallas, TX; Division of Cardiology, University of Texas Southwestern, Dallas, TX; Division of Endocrinology, Department of Internal Medicine, University of Texas Southwestern, Dallas, TX; Institute for Healthcare Policy and Innovation, and Division of Cardiovascular Medicine, Department of Internal Medicine, University of Michigan, Ann Arbor, MI

**Keywords:** familial hypercholesterolemia, hyperlipidemia, disparities, race, ethnicity, sex

## Abstract

**Background:** Individuals with familial hypercholesterolemia (FH) require intensive lipid-lowering therapy, starting with high-intensity statins and adding ezetimibe and PCSK9 inhibitors (PCSK9i) as needed to reach target LDL-C levels. There are limited data on disparities in the use of these therapies among individuals with FH in the US.

**Methods:** We queried a large US healthcare claims repository consisting of 324 million individuals, focusing on prescriptions for high-intensity statins, ezetimibe, and PCSK9i in two patient groups: those diagnosed with FH (ICD-10 E.78.01) and those not diagnosed with FH but identified as having probable FH (PFH) via the FIND-FH® machine learning algorithm. We used multivariable regression models to examine correlations with demographic/socioeconomic variables.

**Results:** In the FH cohort (n = 85,457), 45.9% were female, 79.4% identified as White, 12.2% Black, and 8.4% as Hispanic. In the PFH cohort (n = 287,580), 42.2% were female, 78.2% White, 13.7% as Black, and 8.1% as Hispanic. Males were more likely to be prescribed high-intensity statins than females: odds ratio (OR) [95% confidence interval (CI)] = 2.05 [1.97, 2.13] and 1.60 [1.56,1.63] in the FH and the PFH cohorts, respectively. In both cohorts, White individuals were more likely to get ezetimibe, PCSK9i, or combination therapy compared to Black individuals (ORs: 1.12-1.40). Higher income was associated with increased odds of receiving these treatments (OR: 1.17-1.59 for incomes >$50,000). Higher education was linked to a higher likelihood of combination therapy (ORs [95% CI] = 1.49 [1.33, 1.68] and 1.18 [1.10, 1.27] in the FH and PFH cohorts, respectively).

**Conclusions:** Real-world data indicate that more aggressive lipid-lowering therapy (ezetimibe and PCSK9i) is more often prescribed to White individuals, individuals with higher income, or those with advanced education, highlighting the need to improve equity in cardiovascular risk reduction for all individuals with FH.

**Clinical Perspective:** *What is new?:*

- Health disparities exist for individuals with FH, regardless of whether they’ve been diagnosed.
- The use of ezetimibe and PCSK9i is more likely to occur in individuals who are White, higher income, and advanced education.

*What are the clinical Implications:*

- Addressing health disparities in FH requires a multifaceted approach, including systemic changes to reduce bias in healthcare systems.
- Ensuring equitable access to expensive lipid medications, such as PCSK9 inhibitors, is crucial for individuals from lower socioeconomic groups.
- Providing access to case managers and geneticists can enhance health education and support, ultimately improving treatment outcomes for individuals with FH.

## Introduction

Familial hypercholesterolemia (FH) is an autosomal dominant disorder characterized by severe elevations in low-density lipoprotein cholesterol (LDL-C) from birth. Individuals with FH are at significantly increased risk for atherosclerotic cardiovascular disease (ASCVD), and aggressively reducing LDL-C levels through lipid-lowering drugs can mitigate this escalated ASCVD risk.^1^

The 2018 multi-society cholesterol guidelines recommend first-line treatment with high-intensity statins and, as needed, the addition of ezetimibe and PCSK9 inhibitors (PCSK9i) to achieve an LDL-C target of <100 mg/dL for primary prevention or a <70 mg/dL for secondary prevention.^2^ However, less than half of individuals with FH attain treatment goals. ^3,4^

We previously identified health disparities in LDL-C goal attainment and statin usage from specialized US lipid clinics within the CASCADE-FH® (CAscade SCreening for Awareness and DEtection) registry, a nationwide initiative of the Family Heart Foundation to profile treatment trends and track clinical and patient-reported outcomes in FH patients.^5^ We observed that women were less likely than men to receive high-intensity statin therapy, and non-Hispanic Black (“Black”) individuals were less likely than non-Hispanic White (“White”) individuals to meet LDL-C treatment targets, despite the former being more likely to receive high-intensity statins. However, it remains uncertain whether comparable disparities 1) extend to the wider US FH population, 2) include the use of ezetimibe and PCSK9i, and 3) relate to socioeconomic factors. Examining socioeconomic factors is especially crucial, as misguided beliefs about the biological basis of race have increased health care disparities.

We hypothesized that certain subpopulations, often from marginalized demographic or socioeconomic backgrounds, such as women, non-White individuals, those with limited educational attainment, and lower income levels, may be less likely to receive more aggressive lipid-lowering treatments (i.e., ezetimibe and PCSK9i) crucial for FH management. Such disparities would be concerning due to FH’s impact across all racial and ethnic groups and the elevated risk of premature ASCVD in women with FH, even during their reproductive years.

## Methods

### Study population

For the data included in our analyses, we queried the Family Heart Database™. This database comprises diagnostic, procedural, and prescription data derived from claims and/or laboratory records, encompassing a vast cohort of over 324 million individuals within the United States. The data in the Family Heart Database™ used for our analyses were sourced from Symphony Health (ICON plc Company, Blue Bell, PA), as previously described.^6^ For the current analysis, the dataset spanned from Sept 1, 2016 to Jun 30, 2020 and consisted of more than 77 million individuals with adequate prescription data covering the preceding six years.

Inclusion criteria for this study comprised of individuals meeting either of two conditions: those with a diagnosis of FH, defined by an ICD-10 diagnosis code of E.78.01, or those with undiagnosed FH, referred to as “probable FH” (PFH) as identified by the previously validated FIND FH® machine learning model developed by the Family Heart Foundation.^7^ Of note, that the FIND machine learning model excludes individuals with the ICD-10 E78.01 diagnostic code for FH.

Our analysis focused on examining filled prescriptions for high intensity statins, ezetimibe, and PCSK9i. The patients’ self-identified racial and ethnic classifications encompassed non-Hispanic Black (“Black”), non-Hispanic White (“White”), Hispanic, and the “Other” category. For the analyses described below, we opted to exclude the “Other” category - constituting of 3.30% and 3.25% of the patients in the FH and PFH cohorts, respectively – since this category included multiple ethnic/racial groups, potentially confounding associations between disparities in filled prescriptions and socioeconomic variables.

The dataset provided access to two key socioeconomic factors: education and household income. Education was divided into three levels: “high school graduate or less,” “some college,” and “associate / bachelor’s degree or higher.” Income was classified into three groups: “<$50K,” “$50-100K,” and “>$100K.”

### Lipids lowering therapy categorization

Based on the analysis of filled prescriptions for statins, ezetimibe, and PCSK9i, patients within the LLT group were sorted into four distinct yet non-exclusive categories:

1. High-Intensity statin: filled prescriptions for rosuvastatin at 20 or 40 mg per day or atorvastatin at 40 or 80 mg per day as defined by the 2018 Cholesterol Guidelines.^8^
2. Ezetimibe: filled prescription for ezetimibe.
3. PSCK9i: filled prescriptions for alirocumab or evolocumab administered either biweekly or monthly. (Inclisiran was not included as it was not approved until December 2021.)
4. Statin + Ezetimibe + PCSK9i (“combination therapy”): filled prescriptions for statin along with ezetimibe and PCSK9i.

### Statistical analysis

Data were summarized by count (proportion) for categorical variables and mean ± standard deviation for continuous variables. Comparisons among groups were made by t-test, ANOVA, and Pearson’s chi-squared test as appropriate. Multiple logistic regression models were fit with the binary LLT type as the dependent variable and the demographic and socioeconomic factors—race/ethnicity, age, sex, income, and education— as independent variables. Odds ratio (OR) and 95% confidence interval (CI) were reported. Analyses were performed in all individuals as well as in each racial/ethnic group, separately. We also performed sensitivity analysis in male and female patients, separately, and in three age groups: lower 50 percent, upper 50 percent, and interquartile range. All statistical analysis was performed using R (v4.2.1).

## Results

### Characteristics

Table 1 provides a summary of the demographic characteristics of patients categorized by racial and ethnic classification. There were 85,457 and 287,580 individuals In the FH and PFH cohorts, respectively. There were fewer Black individuals in the FH cohort than in the PFH cohort (12.2% vs. 13.7%; *P*<0.001). There were more females in the FH cohort than in the PFH cohort for all three ethnic groups (*P’*s<0.001). There was no significant difference between the two cohorts regarding education or income for Black and Hispanic groups. In contrast, the White group in the FH cohort tended to have lower education and income levels than those in the PFH group (*P’*s<0.005). The average age of patients in each subgroup was above 60 years old with the White group being the oldest group and Hispanic group the youngest in both cohorts (*P’*s<0.001). In each cohort, the White group displayed the highest levels of education and income (*P’*s<0.001).

**Table 1:** Demographic distribution of familial hypercholesterolemia (FH) and probable familial hypercholesterolemia (PFH) patients receiving lipid lowering therapies stratified by race and ethnicity.

|  | FH |  |  | PFH |  |  |
| --- | --- | --- | --- | --- | --- | --- |
|  | Black | White | Hispanic | Black | White | Hispanic |
| Sample Size | 10405<br>(12.2%) | 67883<br>(79.4%) | 7169<br>(8.4%) | 39317<br>(13.7%) | 225029<br>(78.2%) | 23234<br>(8.1%) |
| Age, years | 62.9 (11.5) | 63.3 (11.7) | 61.6 (12.3) | 63.1 (11.57) | 64.6 (11.3) | 62.0 (12.0) |
| Sex |  |  |  |  |  |  |
| Female | 5755<br>(55.3%) | 29944<br>(44.1%) | 3498<br>(48.8%) | 21003<br>(53.4%) | 89902<br>(40.0%) | 10423<br>(44.9%) |
| Male | 4650<br>(44.7%) | 37939<br>(55.9%) | 3671<br>(51.2%) | 18314<br>(46.6%) | 135127<br>(60.0%) | 12811<br>(55.1%) |
| Education |  |  |  |  |  |  |
| High school graduate or less | 5995<br>(57.6%) | 24849<br>(36.6%) | 3785<br>(52.8%) | 23000<br>(58.5%) | 78688<br>(35.0%) | 12267<br>(52.8%) |
| Some college | 3556<br>(34.2%) | 26631<br>(39.2%) | 2289<br>(31.9%) | 13177<br>(33.5%) | 89598<br>(39.8%) | 7314<br>(31.5%) |
| Associate / bachelor's degree or more | 854 (8.2%) | 16403 (24.2%) | 1095 (15.3%) | 3140 (8.0%) | 56743 (25.2%) | 3653 (15.7%) |
| Income |  |  |  |  |  |  |
| <\$50K | 5852<br>(56.2%) | 20360<br>(30.0%) | 3286<br>(45.8%) | 22119<br>(56.3%) | 66225<br>(29.4%) | 10423<br>(44.9%) |
| \$50-100K | 3351<br>(32.2%) | 25235<br>(37.2%) | 2467<br>(34.4%) | 12934<br>(32.9%) | 83592<br>(37.1%) | 8002<br>(34.4%) |
| >\$100K | 1202<br>(11.6%) | 22288<br>(32.8%) | 1416<br>(19.8%) | 4264<br>(10.8%) | 75212<br>(33.4%) | 4809<br>(20.7%) |

Data are presented as counts (proportion) exception that age is presented as mean (standard deviation). For each variable, comparisons among ethnic groups within a cohort and between cohorts for an ethnic group were performed by t-test, ANOVA and Pearson's chi-squared test as appropriate. Bonferroni correction was applied. Except that there was no significant difference between the two cohorts regarding education or income for Black and Hispanic groups, all other comparisons were statistically significant ( $P$ 's<0.001).

### Differences in filled prescriptions between ethnic/racial groups

Table 2 presents the proportion of individuals across various types of LLT based on race and ethnicity. Figure 1 indicates the association of race and ethnicity with the filled prescription patterns of LLT as determined through multivariable analysis.

**Figure 1:**
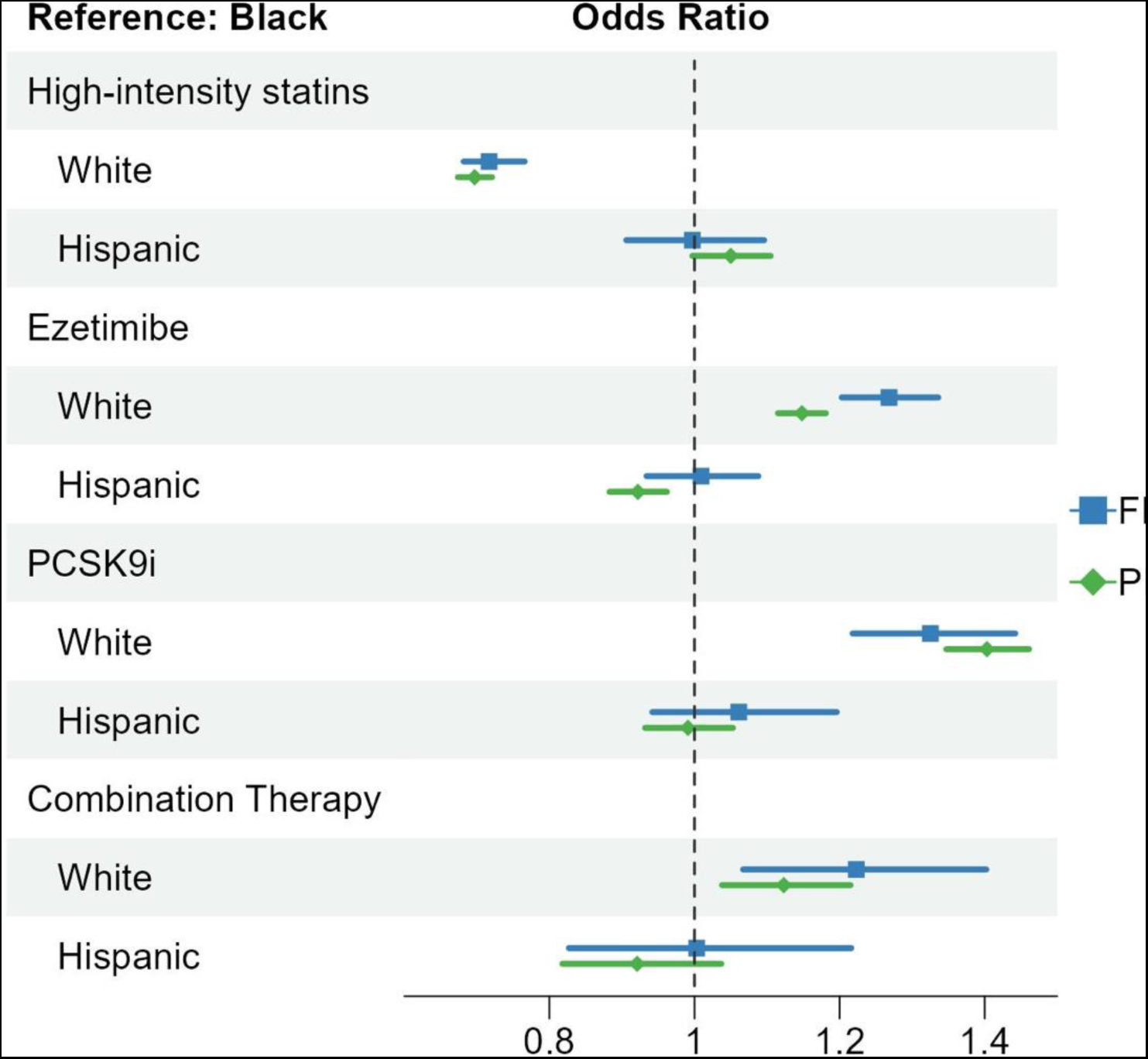
Association of race and ethnicity with filled prescription for lipids lowering therapies. Multiple logistic regression models were fit with the binary lipids lowering therapy type as the dependent variable and ethnicity as the independent variable adjusting for age, sex, income, and education. The reference group was non-Hispanic Black. The FH cohort and PFH cohort were analyzed separately. Sample sizes are reported in Table 2. Odds ratio and 95% confidence interval were plotted.

**Table 2:** Proportions of familial hypercholesterolemia (FH) and probable familial hypercholesterolemia (PFH) patients receiving lipid lowering therapies stratified by race and ethnicity.

|  | FH |  |  | PFH |  |  |
| --- | --- | --- | --- | --- | --- | --- |
|  | Non-Hispanic Black | Non-Hispanic White | Hispanic | Non-Hispanic Black | Non-Hispanic White | Hispanic |
| N | 10405 | 67883 | 7169 | 39317 | 225029 | 23234 |
| High-intensity statin | 9210 (88.5%) | 56985 (83.9%) | 6349 (88.6%) | 34677 (88.2%) | 186306 (82.8%) | 20642 (88.8%) |
| Ezetimibe | 2078 (20.0%) | 17081 (25.2%) | 1469 (20.5%) | 7012 (17.8%) | 47875 (21.3%) | 3928 (16.9%) |
| PCSK9i | 682 (6.6%) | 6130 (9.0%) | 522 (7.3%) | 2993 (7.6%) | 25288 (11.2%) | 1772 (7.6%) |
| Statin + Ezetimibe + PCSK9i | 249 (2.4%) | 2167 (3.2%) | 188 (2.6%) | 767 (2.0%) | 5428 (2.4%) | 441 (1.9%) |

Data are presented as counts (proportion). Statistical comparisons were described in Figures 1-4.

#### High-intensity statins

Within both FH and PFH, White individuals exhibited a decreased likelihood of receiving high-intensity statins when compared to Black individuals: ORs [95% CI] were 0.72 [0.68, 0.77] and 0.70 [0.67, 0.72] in the FH cohort and PFH cohort, respectively. There were no statistically significant distinctions between the Black and Hispanic groups.

#### Ezetimibe, PCSK9i, and combination therapy

In the FH cohort, White individuals were more likely to receive ezetimibe, PCSK9i, and the combination of statin+ezetimibe+PCSK9i treatments—ORs [95% CI] equal to 1.27 [1.20,1.34], 1.33 [1.22,1.44], and 1.22 [1.07,1.40], respectively—compared to Black individuals, whereas there was no statistical difference between Hispanics and Black individuals (Figure 1).

In the PFH cohort, White individuals were more likely to receive ezetimibe, PCSK9i, and the combination of statin+ezetimibe+PCSK9i treatments—ORs [95% CI] equal to 1.15 [1.12,1.18], 1.40 [1.35,1.46], and 1.12 [1.04,1.21], respectively—compared to Black individuals; Hispanic individuals were less likely to receive ezetimibe compared to Black individauls OR [95% CI] = 0.92 [0.88,0.96], but there was no significant difference for PCSK9i or the combination therapy (Figure 1).

### Differences in filled prescriptions among income groups

#### High-intensity statins

As income levels increased, the OR of a filled prescription for high-intensity statins decreased (Figure 2). Relative to the “<$50K” income group, the ORs [95% CI] for receiving high-intensity statins were 0.77 [0.73, 0.81] and 0.61 [0.58, 0.65] for the “$50-100K” and “$>100K” income groups, respectively, within the FH cohort. These associations were consistently observed in the PFH cohort as well.

**Figure 2:**
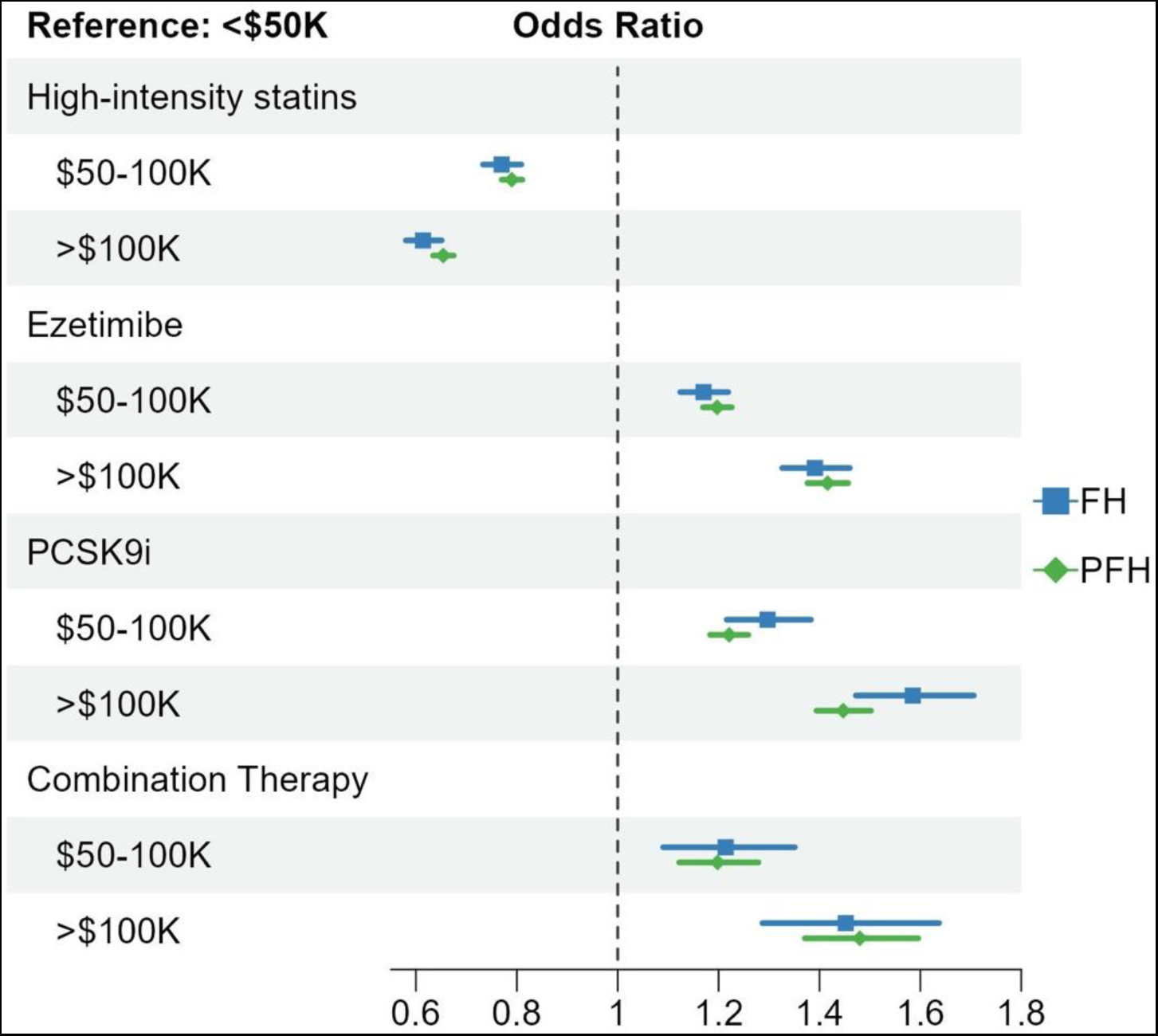
Association of income levels with filled prescription for lipid lowering therapies. Multiple logistic regression models were fit with the binary lipids lowering therapy type as the dependent variable and income as the independent variable adjusting for age, sex, ethnicity, and education. The reference group was yearly income “<$50K”. The FH cohort and PFH cohort were analyzed separately. Sample sizes are reported in Table 2. Odds ratio and 95% confidence interval were plotted.

#### Ezetimibe, PCSK9i, and combination therapy

As income levels increased, the OR of a filled prescription increased for ezetimibe, PCSK9i, and the combination therapy (Figure 2). Compared to the “<$50K” income group, the ORs [95% CI] of receiving the three treatments were 1.17 [1.12,1.22], 1.30 [1.22-1.38], and 1.21 [1.09,1.35], respectively, for the “$50-100K” group, and 1.39 [1.33,1.46], 1.59 [1.47,1.71], and 1.45 [1.29,1.64] for the “>$100K” group in the FH cohort. These associations were consistently observed in the PFH cohort as well.

### Differences in filled prescriptions among education groups

#### High-intensity statins

Higher education levels were linked to a lower likelihood of high-intensity statins prescription in the PFH cohort—Relative to the “high school graduate or less” group, the ORs [95% CI] for receiving high-intensity statins were 0.94 [0.90, 0.99] and 0.93 [0.88, 0.98] for the “some college” and “ associate / bachelor’s degree or more” groups, respectively. These associations were not statistically significant in the PFH cohort (Figure 3).

**Figure 3:**
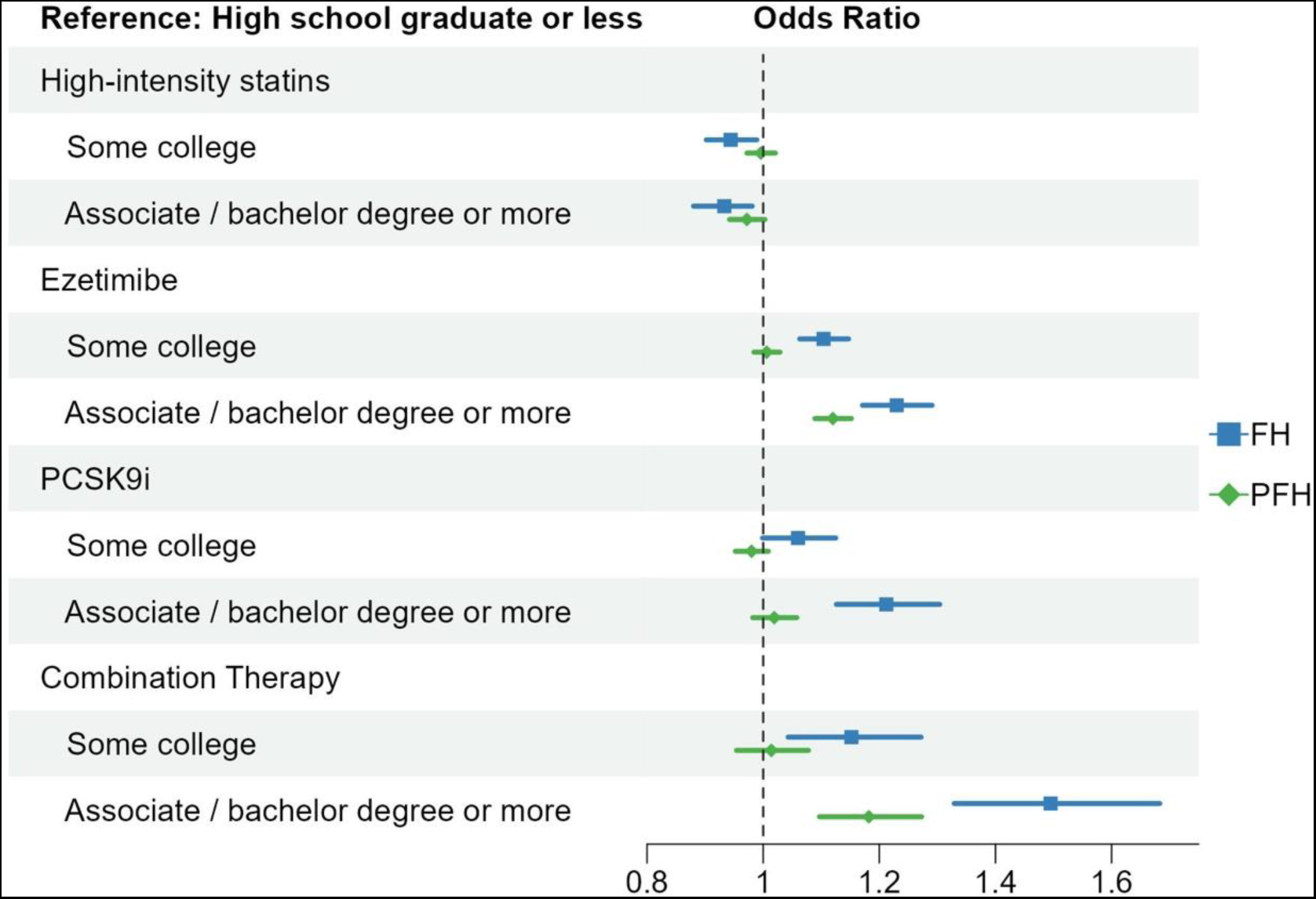
Association of education levels with filled prescription for lipids lowering therapies. Multiple logistic regression models were fit with the binary lipids lowering therapy type as the dependent variable and education as the independent variable adjusting for age, sex, ethnicity, and income. The reference group was education level of “high school graduate or less”. The FH cohort and PFH cohort were analyzed separately. Sample sizes are reported in Table 2. Odds ratio and 95% confidence interval were plotted.

#### Ezetimibe, PCSK9i, and combination therapy

The likelihood of an individual getting a filled prescription for ezetimibe, PCSK9i, and the combination therapy increased with higher education levels. Within the “associate / bachelor’s degree or more” group, the ORs [95% CI] for receiving these treatments were as follows: 1.23 [1.17, 1.29] for ezetimibe, 1.21 [1.13, 1.30] for PCSK9i, and 1.50 [1.33, 1.68] for the combination therapy, relative to the “high school graduate or less” group in the FH cohort. This association remained significant for ezetimibe and the combination therapy in the PFH cohort, with ORs [95% CI] of 1.12 [1.09, 1.15] and 1.18 [1.10, 1.27], respectively.

### Differences in filled prescriptions by gender

Males displayed a higher likelihood of obtaining filled prescriptions for high-intensity statins— ORs [95% CI] were 2.05 [1.97, 2.13] and 1.60 [1.56, 1.63] in the FH cohort and PFH cohort, respectively (Figure 4). In contrast, males exhibited a decreased likelihood of being prescribed ezetimibe, PCSK9i, and the combination therapy, with ORs [95% CI] of 0.65 [0.63, 0.67], 0.57 [0.54, 0.60], and 0.64 [0.69, 0.70], respectively, in the FH cohort, and ORs [95% CI] of 0.76 [0.74, 0.78], 0.83 [0.81, 0.85], and 0.86 [0.82, 0.90], respectively in the PFH cohorts.

**Figure 4:**
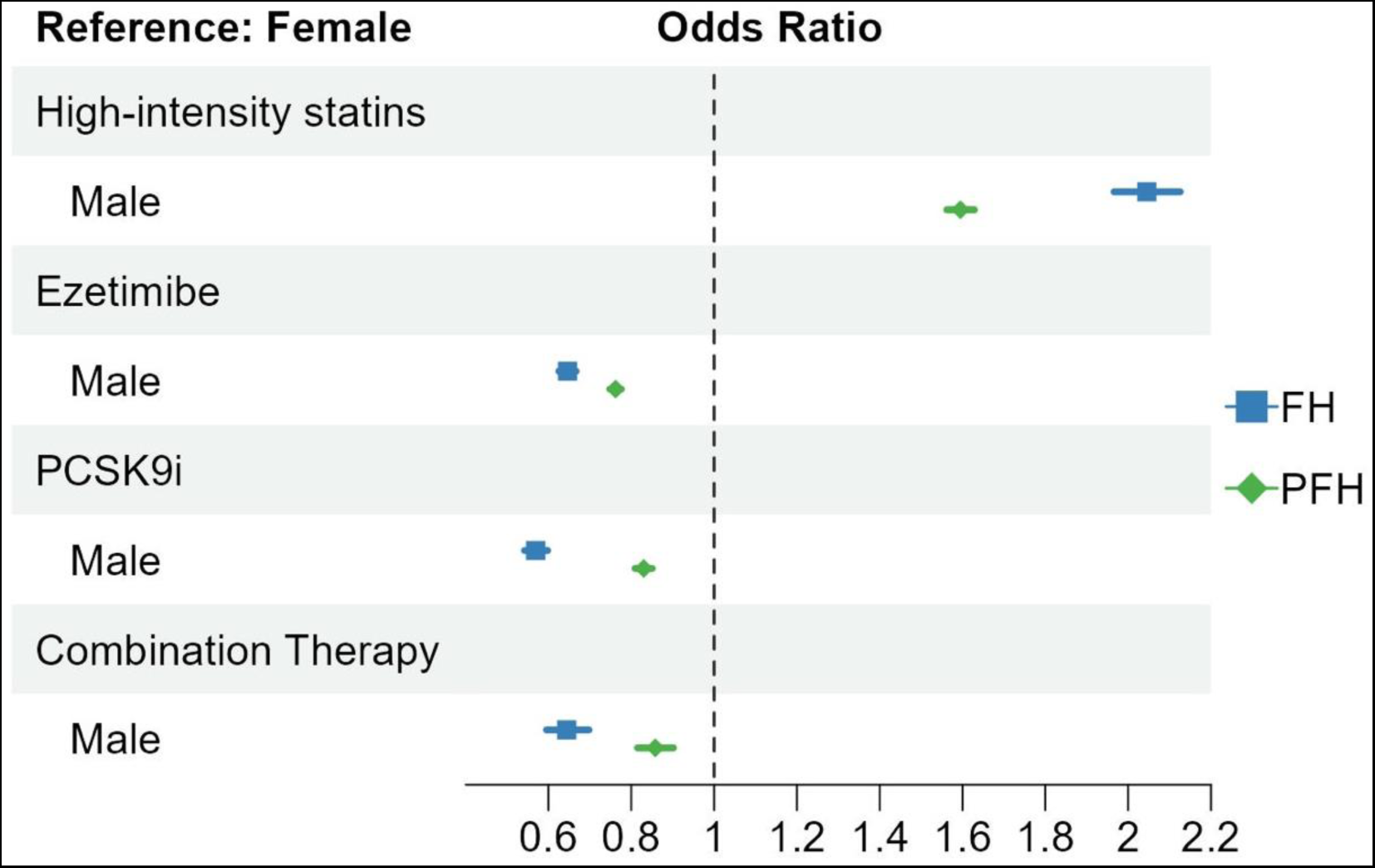
Association of gender with filled prescription for lipids lowering therapies. Multiple logistic regression models were fit with the binary lipids lowering therapy type as the dependent variable and sex as the independent variable adjusting for age, ethnicity, income, and education. The reference group was female. The FH cohort and PFH cohort were analyzed separately. Sample sizes are reported in Table 2. Odds ratio and 95% confidence interval were plotted.

### Sensitivity analyses

Sensitivity analyses were performed in the subgroups stratified by race—White, Black, and Hispanics (Figures S1-S3), gender—males and females (Table S1), and age—lower 50 percent, upper 50 percent, and interquartile range (Table S2). Most of findings using the whole sample remained significant: White individuals were more likely to receive ezetimibe, PCSK9i, and the combination therapy than Black and Hispanic individuals; White individuals were less likely to receive high-intensity statins than Black individuals; males were more likely to receive high-intensity statins; the likelihood of being prescribed ezetimibe, PCSK9i, and the combination therapy all increased with higher education and greater income.

## Discussion

We analyzed a healthcare claims database and found treatment disparities for FH patients in the US population with regards to filled prescription for high-intensity statins, ezetimibe, and PCSK9i. Our main finding is that that more aggressive lipid-lowering therapy (ezetimibe and PCSK9i) is more often prescribed to White individuals, individuals with higher income, or those with advanced education.

Our findings contribute to the existing body of knowledge concerning health disparities among individuals with FH. Prior work has mostly been done outside the US, limited to gender differences, and did not include individuals with FH who have yet to be diagnosed (PFH). In a FH cohort from British Columbia (n = 579), Ryzhaya et. al. also found women were less likely to be prescribed high-intensity statins (45% women vs 55% p=0.03).^9^ Unlike our findings, though, they found females were less likely to be prescribed ezetimibe; in addition, socioeconomic factors were not accounted for in their analyses. In a separate cohort of FH patients in Spain, PCSK9i use was uncommon (3.5%) – similar to our findings – and women were less frequently treated with high-intensity statins (OR [95% CI] = 0.62; 0.44-0.88) and PCSK9 inhibitors (OR [95% CI] = 0.56; 0.39-0.82).^10^

In the US population of individuals with FH, previous research has offered limited exploration of disparities beyond the scope of statin usage, and only a few studies have incorporated socioeconomic factors into their analyses. Although not specific to FH, a recent study from the US Veterans Affairs system identified 1,225 veterans with severe hypercholesterolemia (LDL-C above 190 mg/dL). Similar to our findings in women, their data showed females were more likely to be initiated on PCSK9i (OR 1.99; 1.03-3.87).^11^ However, it’s noteworthy that within this cohort of severe hypercholesterolemia patients in the Veterans Affairs system, factors such as ethnicity, race, and income level were not identified as significant determinants. This observation potentially reflects the specific patient population accessing care through the Veterans Affairs system, where different socioeconomic factors and structural determinants of health might be at play.

In comparison to the disparities observed in the general population, our findings for high-intensity statins differ from established patterns. Typically, research in the general population has revealed that cohorts who are Black or lower socioeconomic status tend to exhibit the lowest utilization of statins^12–14^. However, we previously found, among Black individuals with FH who get care at specialty lipid clinics, a higher chance of getting high-intensity statin therapy. ^5^ Our current study, in the broader US context of individuals with FH, replicates these finding and also identified lower socioeconomic status (income and education) as a predictor of high-intensity statins use. These differences between the general population and the FH population suggest that factors specific to FH may influence prescription patterns, resulting in unique disparities that deviate from broader trends. This emphasizes the importance of conducting targeted research within specific patient populations, such as FH, to gain a comprehensive understanding of the complexities surrounding health disparities and treatment practices.

In the general population, women are less likely to receive guideline-recommended statin therapy compared with men.^15,16^ This observed disparity has been attributed to a variety of factors including refusal of statin therapy, increased discontinuation of statin therapy, and potentially lower prescription rates of statin therapy by their healthcare providers.^15,17–19^ Our study’s findings introduce an additional potential explanation for this phenomenon: women, as a collective, might utilize ezetimibe and PCSK9i more so compared to men. It emphasizes the need for further investigation into the factors influencing these prescription patterns to ensure equitable and optimal cardiovascular care for all individuals.

Our findings of disparities associated with income levels are especially important since income is strongly associated with morbidity and mortality, and income-related health disparities appear to be growing. A systematic review and meta-analysis of 12 prospective cohort studies including 1,710,000 participants found that individuals with the lowest income had a 29% higher risk of cardiovascular disease compared to those with the highest income. Additionally, individuals with low income had a 46% higher risk of dying from cardiovascular disease compared to those with high income. The study concluded that low income is a significant risk factor for cardiovascular disease and related mortality.^20^ The mechanisms underlying the associations between income and education disparities and cardiovascular morbidity and mortality are complex and multifactorial. Social determinants of health, such as education, housing, employment, access to healthcare, and food insecurity are all interconnected and can influence health outcomes, including cardiovascular disease.^21,22^

## Limitations

Despite the robustness of our study’s large sample size, several limitations warrant consideration. Notably, we were unable to account for statin intolerance, a plausible contributing factor in explaining the lower utilization of high-intensity statin therapy in women. Our dataset does not account for differences in ASCVD status. However, it is worth mentioning that prior research encompassing both statin intolerance and ASCVD status found negligible impact on similar overall findings for gender disparities.^23^ Furthermore, certain US healthcare systems, such as the Veterans Affairs and Kaiser Permanente systems, were not encompassed in our dataset. It is important to acknowledge that disparities in health treatment have been identified within these systems as well.^11^ Another potential limitation lies in the possibility of miscoding and errors in provider charting, which may have influenced our results. Our study lacked data pertaining to emerging lipid-lowering medications like inclisiran and bempedoic acid as well as laboratory findings. The absence of information on LDL-C levels restricts our capacity to comment on the achievement of LDL-C goal attainment. Lastly, it is worth noting that our dataset provided access to only a limited set of socioeconomic factors and did not encompass broader structural determinants of health which might play a significant role in health disparities. As such, our data cannot distinguish where these disparities occur in the life experience of FH and PFH patients. One cautionary note on interpreting the results is that the study is subject to collider bias, and therefore one should take caution inferring causality, though the correlations are robust, and the estimates could be biased.

## Impact

As treatment of individuals with FH expands in the US, the disparities we identified may become more pronounced if not addressed. ^24^ Such gaps in lipid treatment have been associated with increased ASCVD morbidity and mortality - as well as increased healthcare expenditures.^25,26^ - especially affecting women, racial and ethnic minorities, and individuals of lower socioeconomic status. ^5,13,27–29^

## Future work

Efforts to improve health disparities must take a multifaceted approach, including addressing systemic discrimination, ensuring equitable access to costly lipid medications, and providing access to case managers and geneticists for health education. Such efforts must consider more socioeconomic factors as well as social determinants of health.^30,31^

## Conclusion/Summary

Real world patterns of medical care reveal lipid lowering therapies, particularly those extending beyond statins to include ezetimibe and PCSK9i, are more often prescribed for individuals with FH who are White, have high income, or received advanced education. Efforts are warranted to improve equity and provide all individuals with FH an opportunity for cardiovascular risk reduction.

## Data Availability

Data available by request, subject to approval by the Family Heart Foundation.

